# Post-Discharge Experiences of Survivors Following the 2022 Ebola Virus Disease Outbreak in Uganda: An Exploratory Qualitative Study

**DOI:** 10.64898/2026.08.29.26361698

**Authors:** Julian Natukunda, Patience Muwanguzi, Tom Denis Ngabirano, Bridget Atuhaire, Sarah Joselyn Nalubega, Christine Auma, Racheal Nabunya

## Abstract

**Background:** Ebola virus disease is a life-threatening illness caused by the Ebolavirus, with symptoms manifesting two to twenty-one days after infection. Although Uganda has faced multiple Ebola outbreaks, many patients survive only to encounter persistent challenges. Therefore, this study explored the post-discharge experiences of survivors following the 2022 Ebola Virus Disease outbreak in Uganda.

**Methods:** An exploratory qualitative study comprising of in-depth participant interviews was conducted at Mubende Regional Referral Hospital in central Uganda. Interviews were face-to-face and data were analyzed manually by inductive content analysis. Ten male and female participants were Ebola Virus Disease survivors in Mubende district who had lived in the community for at least six months post-discharge from the Ebola Treatment Unit.

**Results:** Four themes emerged: (i) Psychosocial Burdens and Social Exclusion, (ii) Economic Hardship and Loss of Financial Stability, (iii) Chronic Physical and Health Burdens Post-Recovery and (iv) Rebuilding Lives: Psychological, Social, and Medical Pathways to Recovery. Survivors faced significant emotional burdens such as survivor guilt, grief, trauma from loss, and anxiety about transmission risks. They experienced social isolation, stigma, and discrimination, which often led to their exclusion from community activities. Financially, they struggled with debt and the loss of livelihoods, compounded by ongoing health issues. Additionally, survivors endured chronic physical complications, including pain and fatigue, which hindered their recovery.

Despite these challenges, survivors sought psychological, social, and medical pathways to recovery, including confirmation of their recovery, support from family and organizations, and health maintenance practices. Supportive medical care and community assistance were crucial in their physical and emotional rehabilitation.

**Conclusion:** Ebola Virus Disease survivors in Uganda face significant psychosocial, health, social, and economic challenges post-discharge. The findings highlight the critical need for comprehensive medical and community-based support systems to aid survivors’ recovery and well-being. Further research on long-term neurological effects and community reintegration programmes is needed to inform targeted interventions that support Ebola survivors and reduce stigma and discrimination.

## Background

Ebola virus disease (EVD) is a severe and acute sickness caused by Ebolavirus (EBOV). The signs of acute disease usually appear two to twenty-one days after infection and include fever, exhaustion, muscle soreness, vomiting, diarrhea, rash, and internal and external bleeding (1). Outbreaks of Ebola disease (EBOD) are a result of both zoonotic and human-to-human transmission by contact with contaminated food or bodily fluids(2, 3). Three species, Zaire ebolavirus, Bundibugyo ebolavirus, and Sudan virus, are known to cause epidemics in Africa, resulting in severe hemorrhagic disease with high case fatality rates (CFR)(4), Zaire ebolavirus has an average case fatality rate (CFR) of 60-90%, while the Sudan virus has a CFR of 40-60%, the Bundibugyo ebolavirus has caused only one outbreak to date with a CFR of 25% (2). Several Ebola outbreaks have been registered in Africa in the last three decades. Uganda has registered 5 outbreaks since 2000, with the largest being the Gulu Outbreak in 2000 (5) .On the 20^th^ of September 2022, the Ministry of Health Uganda confirmed an outbreak of Ebola virus disease caused by the Sudan strain registering 142 confirmed cases, 22 probable cases, 55 confirmed deaths and 87 patients recovered (6, 7).

Despite the high morbidity and mortality of EVD, a high number of survivors has usually been reported. For example, during the 2014-2016 West Africa Ebola outbreak, 35% of the recorded 28,646 cases survived, while in 2000, during the Gulu outbreak in Uganda, 47% of the 425 clinical cases also survived (2), More recently, in the 2022 outbreak in Uganda, 87 out of 142 cases recovered. (7). EVD survivors are known to suffer from both short- and long-term physical symptoms and mental complications such as social stigma (8), depression (9), anxiety and post-traumatic stress disorder (10, 11). In Uganda, misconceptions about Ebola virus disease, influenced by cultural beliefs, contribute to stigma against survivors (12) . Even after recovery, survivors often face social isolation, as community members may fear potential transmission. This stigma significantly impacts their mental health, leading to feelings of fear, shame, depression, and anxiety (13).. Physical role limitation (14) associated with the multisystem involvement has been reported to be widespread, contributing to reduced productivity and quality of life among Ebola survivors (15).

Additionally, EVD survivors face persistent challenges at the community level, including experiences of isolation, rejection, and discrimination (16). It is observed that, despite these difficulties, the generosity of community members expressed through both tangible and intangible support help survivors and those affected by EVD develop a stronger bond with their communities, even in the face of isolation (17).

The predominant focus of studies related to Ebola has been on the biomedical aspects of the illness, including clinical and epidemiological dimensions such as disease transmission, pathogenesis, treatment modalities, and outbreak management (18–20). Additionally, while a few studies have explored the post-discharge experiences of Ebola survivors in other regions, Uganda remains significantly underrepresented in this research—despite experiencing multiple major outbreaks. This gap may stem from the country’s unique epidemiological profile and the distinct cultural, social, and healthcare dynamics that shape survivors’ experiences. Without a thorough understanding of these local contexts, there is a risk that support systems may be inadequate or poorly aligned with survivors’ actual needs. Therefore, this study aimed to explore post-discharge experiences of Ebola survivors in Uganda.

## Methods

### Researcher Characteristics and reflexivity

One co-author, a nurse with first-hand experience working in an Ebola Treatment Unit (ETU), brought valuable insight into the survivors’ challenges, which may have influenced the research approach and interpretation of data. This experience likely shaped the team’s understanding of the emotional, physical, and social difficulties—including stigma—faced by survivors. The other co-authors, also nurses with varying clinical experience, contributed perspectives that may have influenced the study design, particularly in addressing health and psychosocial concerns. Their professional background informed the study’s ethical approach and ensured sensitivity when engaging with trauma-affected participants.

The researchers’ professional roles and prior experiences likely shaped their assumptions and influenced the research focus, particularly on post-discharge experiences, health impacts, and stigma. These presuppositions may have affected how survivors’ accounts were interpreted. While the researchers’ healthcare backgrounds may have fostered trust and encouraged open discussion of sensitive issues, they also introduced the potential for bias in interpreting health-related experiences. Nonetheless, the researchers’ credibility in nursing and healthcare may have strengthened rapport with participants and enhanced the depth and relevance of the data collected, thereby improving the transferability of findings.

### Study design

This study utilized an exploratory descriptive qualitative design(21) within the pragmatic paradigm. Pragmatism offers an action-oriented framework for research, where the researcher aims to tackle practical issues that arise directly from communities by utilizing the most suitable methods to address the research question (22). Therefore, data collection required a flexible approach due to the stigma experienced by the survivors. Some participants were reluctant to allow additional health vehicles to visit their homes. Furthermore, the sensitivity of the study topic necessitated extended interview periods, with some interviews conducted over multiple sessions to accommodate the resurfacing of traumatic memories. This flexibility is a hallmark of the pragmatic approach, which prioritizes the needs of participants to ensure meaningful data collection.

### Study Setting and Participants

This exploratory study was conducted in April 2023 at Mubende Regional Referral Hospital, situated in the central region of Uganda. During the 2022–2023 Ebola Virus Disease (EVD) outbreak, Mubende became the epicenter, accounting for 45% of the confirmed cases, with 64 confirmed and 19 probable cases. To address the outbreak, an Ebola Treatment Unit (ETU) was established at the hospital to provide specialized care for Ebola patients (7). This government-owned health facility operates under the national Ebola Survivors’ Programme, which is dedicated to delivering high-quality medical care, mental health, and psychosocial support services to Ebola survivors. Through collaboration with various health implementation partners, the program aims to support survivors in achieving full recovery (23). Ten Ebola Virus Disease (EVD) survivors—both male and female—who had lived in the community for at least six months post-discharge, were enrolled through the Survivors’ Unit and consented to participate in the study. Survivors with health or psychological limitations that hindered their ability to provide reliable information—such as memory loss, hearing impairment, or significant emotional distress—were excluded.

### Data collection

Permission to access records for identifying eligible participants was obtained from the survivors’ clinic at Mubende Regional Referral Hospital. Prior to data collection, 15 eligible Ebola virus disease survivors were approached using purposive sampling. Of these, 10 provided informed consent and participated in the study. The remaining five declined, citing emotional discomfort and a reluctance to revisit distressing experiences related to their illness and post-discharge life. Their decisions were respected, and no further attempts were made to persuade them. Ten (10) participants were purposively sampled to include five male and five female individuals who had contracted and recovered from EVD.

Data collection was conducted through in-depth interviews by two trained team members with experience in qualitative research. A structured interview guide, developed by the principal investigator, incorporated open-ended questions informed by a review of existing literature(24, 25) and consultations with Ebola care experts (Supplementary file 1). The guide was pilot tested with two participants to refine the questions, ensure comprehensive coverage of the necessary topics, and maintain appropriate length and flow. The pilot participants provided written informed consent before participating.

Interviews were conducted face-to-face in private, convenient locations chosen by the participants. Each session lasted approximately 40 minutes to one hour and was audio-recorded with the participants’ prior consent. The interviews explored the lived experiences of EVD survivors. The study population of Ebola survivors is rare and often faces significant stigma within their communities, making recruitment challenging. Our goal was to obtain rich and meaningful narratives from the 10 participants who consented to take part in the study. All participants provided written informed consent, including their agreement to have the interviews audio recorded. During data collection, participants who exhibited emotional distress related to their experiences with Ebola disease were referred to the research team’s psychologist for additional psychosocial support. Those requiring medication or more specialised mental health care were further referred to a psychiatrist at the regional referral hospital.

### Data analysis

The audio recordings and transcripts were securely stored to ensure participant confidentiality. Audio files were saved on a password-protected computer accessible only to the research team, and transcripts were stored in a locked location. To maintain anonymity, each participant was assigned a unique identification number. Audio recordings were transcribed verbatim to preserve the accuracy of the data.

Two groups of researchers (JN, BA, and RN, JSN) independently reviewed the transcripts for accuracy and interpretation. Each group generated initial codes, which were then discussed to reach a consensus. Any disagreements were resolved through further deliberation with additional team members, including PM. Qualitative data analysis was conducted using the inductive content analysis method, allowing themes to emerge naturally from the data (26). Data with similar patterns or recurring themes was systematically organized and condensed into groups, from which codes were generated. These codes were further categorized and refined into sub-themes. To provide authenticity and context, direct quotations from study participants were included and presented in italics.

### Trustworthiness

The data underwent repeated review and cross-referencing with the interview guide notes to ensure all responses were accurately captured and interpreted during analysis. To enhance trustworthiness and credibility, two participants—one male and one female—were invited to review the identified themes and sub-themes. Their feedback confirmed that the findings accurately reflected their experiences and responses.

### Ethical consideration

Approval to conduct the study was obtained from the Makerere University School of Health Sciences Research and Ethics Committee (MAKSHSREC-2023-497). Institutional permission to carry out the study was sought from the Head of the Ebola Survivors’ Unit at Mubende Regional Referral Hospital. A written informed consent including an agreement to have the interviews audio recorded, was provided by all enrolled participants. Participants did not receive any financial compensation, as interviews were conducted during their routine review visits to the Survivors’ Treatment Unit. This was a self-sponsored student-led study. All participants were clearly informed that participation was entirely voluntary, not part of their medical care, and that they could withdraw from the study at any point without any consequences to their treatment.

### Patient and Public Involvement statement

Patients or the public were not involved in the design, or conduct, or reporting, or dissemination plans of our research.

## Results

### Participants’ Characteristics

In total, ten (10) Ebola Virus Disease survivors were interviewed. The majority of the participants were married (60%). Half of the participants were aged 30-40 years (50%) Most participants had either no formal education (40%) or only completed primary school (40%), while only 20% attained tertiary education (Table 1).

**Table 1:**
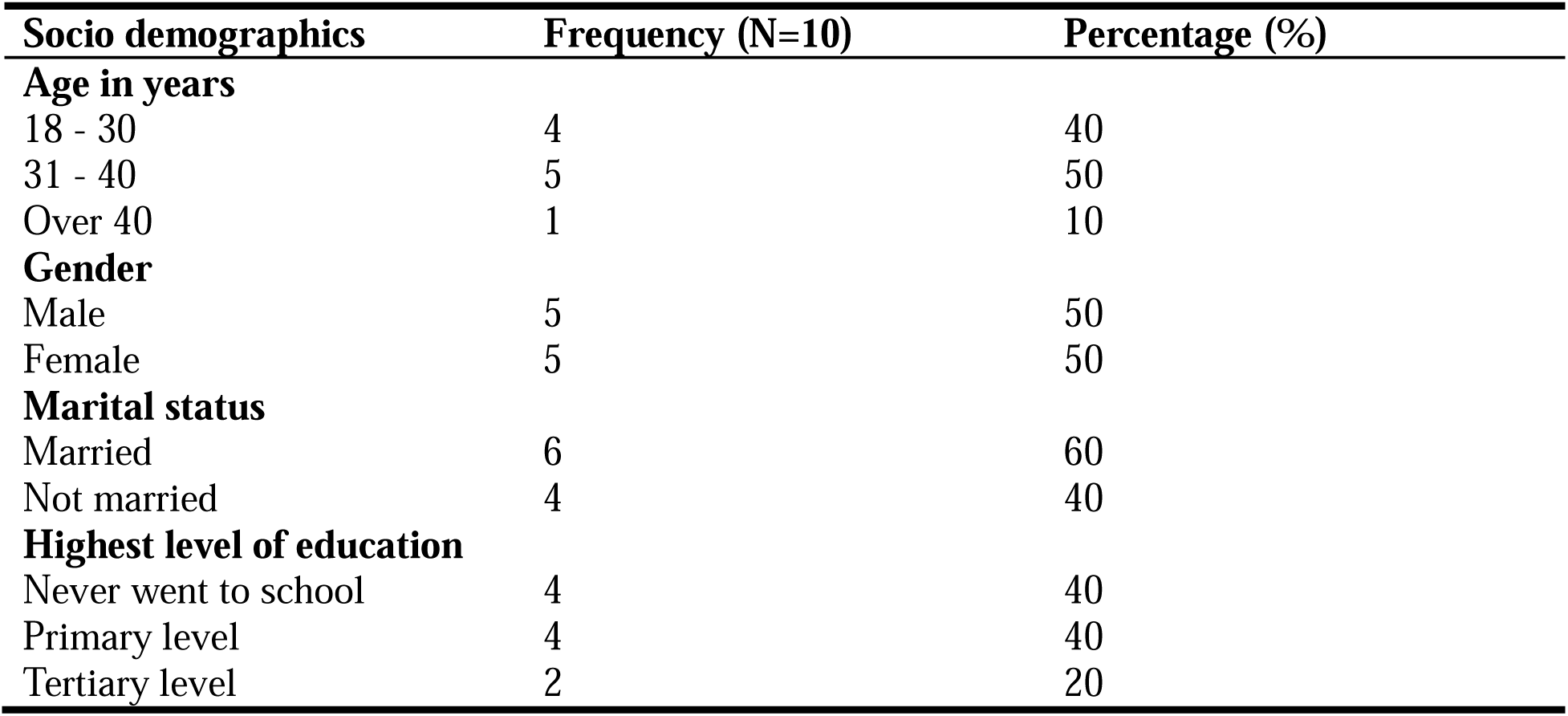
Socio-demographic characteristics of Ebola virus disease survivors, post discharge from Mubende Regional Referral Hospital (N=10)

| Socio demographics | Frequency (N=10) | Percentage (%) |
| --- | --- | --- |
| **Age in years** |  |  |
| 18 - 30 | 4 | 40 |
| 31 - 40 | 5 | 50 |
| Over 40 | 1 | 10 |
| **Gender** |  |  |
| Male | 5 | 50 |
| Female | 5 | 50 |
| **Marital status** |  |  |
| Married | 6 | 60 |
| Not married | 4 | 40 |
| **Highest level of education** |  |  |
| Never went to school | 4 | 40 |
| Primary level | 4 | 40 |
| Tertiary level | 2 | 20 |

### Lived experiences of Ebola virus disease survivors post discharge from Ebola treatment unit in Uganda

The lived experiences are presented under four themes.

### Psychosocial Burdens and Social Exclusion

This theme captures the profound emotional and social challenges faced by Ebola Virus Disease survivors in Uganda after their discharge. It encompasses the deep feelings of survivor guilt, grief, and trauma from the loss of loved ones, as well as the intense anxiety and fear of transmitting the disease. Additionally, the theme highlights the survivors’ experiences of social isolation, displacement, and stigma, reflecting the ongoing psychological and social struggles they face as they navigate life after recovery.

#### Survivor Guilt and Profound Grief

Survivors expressed guilt for perceived roles in spreading the disease and the resulting loss of lives. This sense of responsibility for spreading illness led many to blame themselves for the resulting loss of life. Additionally profound grief emerged as participants mourned the deaths of family, friends, and healthcare providers. Others witnessed the deaths of their loved ones, which was a traumatic experience that continues to affect them. These survivors expressed their pain through tears and noticeable mood changes.

> *“I also blame myself for most of the people that died here because like my neighbour (local healer) here who we went with us pick the child, I got her from the garden to help me with the treatment of my brother’s child and she died, the child also died, the motorist who brought them died and it was really scary. I wish that I hadn’t called her that day. Even some doctors who attended to us died”* (participant 9, early 30s)

#### Anxiety and Fear of Separation and Transmission

Most participants reported intense anxiety upon diagnosis, fearing for their survival, their children’s futures, and their livelihoods. The high mortality rates deepened their fears, especially for those with young dependents or as sole breadwinners. Quarantine restrictions intensified their fears about their families and uncertain futures, particularly the risk of infecting loved ones. One breastfeeding participant expressed anxiety about transmitting the disease to her son through breastmilk or close contact.

> *“When they told me I had Ebola, what made me saddest and most scared was leaving my child behind. I had to go to the hospital alone since he wasn’t sick, leaving behind a child who was supposed to be breastfeeding. The entire time at the hospital, all I could think about was my son. Now that I’m home, I’m afraid to breastfeed him or even get too close, worried I might infect him.”* (participant 8, late 30s)

#### Social Isolation and Displacement Post-Hospitalization

Some survivors reported feeling abandoned by family, friends, and spouses during their hospitalization due to fears of contagion. As a result, upon discharge, they were forced to seek new places to live, as their family members were not yet ready to welcome them back home.

> *“My mom feared me, and she told me, my son, maybe you haven’t healed well, and you see your grandparents here are old, so it’s better you be in town. She feared, and then I had to return to town”* (participant 6, early 40s)

#### Stigma and community exclusion

Most participants reported experiencing stigma from neighbours and the wider community. They faced discrimination during social events, such as funerals, where they were often labelled as Ebola survivors, even months after being discharged. Exclusion from community activities and avoidance in personal interactions were common, largely due to a lack of public knowledge about the disease. This knowledge gap fostered fear, stigma, and discrimination, contributing to the social isolation of survivors.

> *“And someday I wanted to get some honey, so I called my friend to get me some honey from the supermarket. When he brought it, he stopped at some butcher that is close to my place and called me to find him there. On reaching, he put it at the stall and then he moved back so I could pick it without any contact with him. though I had healed.”* (participant 6, early 40s)

### Economic Hardship

This theme captures the financial challenges experienced by Ebola Virus Disease survivors, including the strain of accumulated debts, increased costs, and the inability to regain livelihoods. It underscores how stigma, ongoing health issues, and the economic impact of hospitalization continue to hinder their ability to achieve financial stability and meet basic needs post-discharge.

#### Job loss and financial instability

Hospitalization led to debts and increased financial burdens, with survivors struggling to meet basic needs post-discharge. In addition, prolonged absence from work led to permanent job loss.

> *“The landlord increased rent because they thought we received money when discharged.”* (participant 3, mid 20s)

#### Health-Related Barriers to Economic Reintegration

Survivors faced difficulties returning to work or regaining financial stability due to stigma and health-related challenges.

> “*When I returned home from the hospital, I found that my shop had been closed. The worker had fled upon learning that I had been diagnosed with Ebola. I reopened the shop, but customers were too afraid to buy anything, fearing they might contract the virus. This continued for several months, and eventually, I had to close the shop because I was paying rent but not making any sales. Now, I am at home with no other source of income, relying on community support and handouts. However, I remain hopeful that things will improve soon*.” (Participant 9, early 30s)

### Chronic Physical and Health Burdens Post-Recovery

This theme captures the persistent physical limitations and ongoing health complications experienced by Ebola Virus Disease survivors in Uganda. It underscores the enduring nature of pain, fatigue, and medical issues that hinder survivors’ recovery and reintegration into everyday life.

#### Impaired Physical Functioning

Many survivors experienced lasting pain, fatigue, and other physical limitations that hindered their post-recovery lives.

> *“Maybe like legs paining and also taking long to get sleep at night. In order to sleep I needed to have worked a lot in the day so that am really overtired and sleep.”* (participant 6, early 40s)

#### Ongoing Health Complications

Survivors reported ongoing medical issues, including pain and discomfort in specific body areas.

> *“That’s when I felt like I needed to be supported. I was experiencing scrotal kind of pain. Then it would come, and it would go like that”* (participant 6, early 40s)

### Rebuilding Lives: Psychological, Social, and Medical Pathways to Recovery

This theme captures the multifaceted aspects of post-discharge recovery for Ebola Virus Disease survivors. It emphasizes the critical interplay between psychological healing, social and economic support, health maintenance, and the essential role of medical care in facilitating survivors’ return to stability and well-being.

#### Personal healing and confirmation of recovery

Six participants expressed profound relief and gratitude upon receiving confirmation that they no longer had the disease, highlighting the importance of psychological healing in their lives.

> *“I was healed. I got the results after they had proved I did not have the disease and I am very happy”* (Participant 1, early 20s)

#### Social and Economic Support Networks

Many survivors benefited from financial and food support from friends, family, and organizations, which helped alleviate their immediate needs and contributed to their efforts to regain stability.

> *“On discharge, they gave me a new mattress, new cups and plates and clothes”* (participant 6, early 40s)
>
> *“I got some money and food from some organization”* (participant 2, mid 20s).

#### Health Maintenance and Adaptive Practices

Most survivors reported taking intentional actions to maintain and improve their physical, mental, and emotional health as part of their recovery.

*“I have started on my pressure medication ever since I got cured” (*Participant 1, early 20s)

*“And now that I recovered, I follow every advice the doctor tells me and I even eat fruits and vegetables” (participant* 2, mid 20s)

#### Supportive Medical Care and Community-Based Assistance

A participant emphasized the exceptional care provided by village doctors during and after their hospital stay, demonstrating the crucial role of medical personnel in the recovery process.

> *“And the doctors in the village really helped me a lot, because there’s even a doctor who used to come and cook for me, food, porridge, and by the time I wake up, everything is there.”* (Participant 1, early 20s)

## Discussion

Four themes emerged: (i) Psychosocial Burdens and Social Exclusion, (ii) Economic Hardship and Loss of Financial Stability, (iii) Chronic Physical and Health Burdens Post-Recovery and (iv) Rebuilding Lives: Psychological, Social, and Medical Pathways to Recovery.

Poor emotional state was one of the key findings among the survivors including guilt, abandonment, loss, grief, depression, anxiety, stigma and discrimination. These symptoms were often attributed to being labelled as “Ebola” and “survivor.” The findings are consistent with those in Congo, Liberia and Sierra Leone (8, 25, 27–30). These further deteriorated the lives of the survivors, led to marriage breakdown as fear-stricken husbands forced wives to return to their parents’ homes (28). Similarly, Gashugi et al. conducted a study on the mental health impact of previous infectious outbreaks, such as Marburg Viral Disease, which is similar to the Ebola outbreak, and found that survivors experience heavy psychological burdens that can be long-lasting (31) Cultural beliefs and misconceptions (32) surrounding Ebola in Uganda plays a significant role in exacerbating these emotional challenges as many individuals in Ugandan communities hold misconceptions about the disease, believing that survivors are still contagious or are somehow cursed (33) The long-term psychological impacts necessitate ongoing mental health interventions, as highlighted by Collier et al., who advocated for community-based mental health support systems (34).

Economic hardships are prevalent among EVD survivors, who often struggle to regain their livelihoods post-recovery. Their poor health led to job rejection and an inability to participate in farming, resulting in decreased household income and food insecurity, which further contributed to their low quality of life. Djomaleu ML et al. noted that the economic reintegration of survivors is often hampered by their health complications and the stigma they face (35) . Additionally, the breakdown of health systems during the outbreak left many survivors without adequate support or resources to rebuild their lives (36). The economic challenges are compounded by chronic health issues, including musculoskeletal pain, ocular complications, and neurological disorders, which can further limit survivors’ ability to work (37, 38). Additionally, survivors experienced a decline in bodily functions, including fatigue, weakness, headaches, vision problems, hearing deficits, and joint pain, which diminished their quality of life and overall wellness. These signs and symptoms are comparable to those reported by survivors in the Democratic Republic of Congo(39). This post-Ebola syndrome is linked to ongoing immune response mechanisms (14, 40, 41). Therefore, it underscores the importance of early treatment to alleviate the symptom burden among survivors, along with the necessity for regular medical follow-ups.

Community engagement and education are essential for fostering resilience among survivors (42). Initiatives aimed at reducing stigma and promoting understanding of EVD can significantly enhance the reintegration process (12, 43). For example, community behavioral change such as communication strategies have been recommended to inform families about the importance of supporting survivors and addressing their unique psychological needs (44). Moreover, survivor networks have been identified as crucial for providing emotional support and facilitating recovery (45, 46). These networks not only help survivors cope with their experiences but also empower them to educate others about the disease, thereby transforming their roles from victims to advocates within their communities.

Economic factors also play a significant role in the rebuilding of lives post-Ebola. Many survivors face economic hardships due to the loss of employment during their illness and the lingering effects of post-Ebola syndrome, which can include physical symptoms such as joint pain and fatigue (47, 48). Addressing these economic challenges is vital for the overall recovery of survivors. Programs that provide vocational training and financial support can help survivors regain their independence and improve their quality of life (47, 48). The integration of psychosocial support with economic assistance is essential for fostering resilience and ensuring that survivors can rebuild their lives effectively. In conclusion, the resilience and rebuilding of lives among Ebola survivors require a comprehensive approach that addresses psychological, social, and economic dimensions. By fostering community support, reducing stigma, and providing economic opportunities, we can help survivors navigate their recovery journey and reintegrate into society successfully.

Further research about long-term neurological effects and effectiveness of community reintegration programs is needed to develop and implement targeted interventions that facilitate the successful community reintegration of Ebola virus disease survivors, while effectively addressing and reducing stigma and discrimination within affected communities.

This study is the first of its kind in Uganda, addressing a significant gap in the literature and offering valuable insights to improve empathetic and effective healthcare and support services. One key limitation was potential research bias, as the co-authors—nurses with first-hand experience in Ebola Treatment Units—may have unintentionally influenced the research process and interpretation. The study’s small sample size also limits the generalizability of the findings. A major challenge encountered was the emotional trauma participants experienced when recounting their post-discharge experiences. Several interviews had to be paused to allow participants to process their emotions. To ensure their well-being, a psychologist on the research team provided immediate psychosocial support, and referrals were made to the regional hospital’s mental health services for participants needing continued care.

## Conclusions and Policy implications

In conclusion, the post-discharge experiences of Ebola Virus Disease survivors in Uganda are marked by significant psychosocial burdens, ongoing health challenges, and profound social and economic impacts. Survivors grapple with survivor guilt, grief, and trauma from loss, while also facing anxiety, fear of transmission, and stigma that contribute to their social isolation. The economic strain, compounded by health-related challenges, further exacerbates their difficulties in rebuilding their lives. The study highlights the need for integrated, multisectoral support mechanisms to aid Ebola survivors in their post-recovery transition. Key policy priorities include the establishment of government-led financial assistance schemes, integration of mental health and psychosocial support services within the primary healthcare system, and the implementation of community-based anti-stigma interventions. These coordinated efforts are essential for promoting survivor reintegration, improving mental health outcomes, and ensuring long-term resilience at both individual and community levels

## Data Availability

The datasets used and analysed during the study are available from the corresponding author on reasonable request.

## List of Abbreviations

CFR: Case fatality rates
EBOV: Ebolavirus
EBOD: Ebola disease
ETU: Ebola Treatment Unit
EVD: Ebola virus disease

## Declarations

### Ethics approval and consent to participate

Ethical review board approval was granted by the Makerere University School of Health Sciences Research and Ethics Committee (MAKSHSREC-2023-497). Institutional permission to carry out the study was sought from the Head of the Ebola Survivors’ Unit at Mubende Regional Referral Hospital. All participants enrolled in the study were adults. No participant under the age of 16 was included; therefore, parental or legal guardian consent was not applicable. Written informed consent, including consent for audio recording of the interviews, was obtained directly from all participants before data collection. The study was conducted in accordance with the Declaration of Helsinki and the Uganda ethical standards.

### Consent for publication

Not applicable.

## Acknowledgements

The authors thank all Ebola disease survivors who participated in this study for generously sharing their experiences. We appreciate the support of healthcare workers and facility administrators during the data collection process. We also acknowledge the College of Health Sciences, Makerere University, and the African Center for Health Equity Research and Innovation (ACHERI) for their institutional support.

## Author Contributions

JN, BA, SJN and RN contributed to the design of the work and data interpretation as well as data collection. PM, TDN and JN contributed to survey design and management of data collection as well as literature review. BA, JN, SJN, CA and RN analysed the data, developed the structure of the article, and wrote the first draft. TDN provided overall supervision. All authors have participated sufficiently in the work to take responsibility for the content, including participation in the conception or design of the work, or the acquisition, analysis or interpretation of data and drafting the work. All authors read and approved the final manuscript.

